# Prediction of OncotypeDX recurrence score using H&E stained WSI images

**DOI:** 10.1101/2025.07.21.25331907

**Authors:** Shachar Cohen, Gil Shamai, Edmond Sabo, Alexandra Cretu, Iris Barshak, Tal Goldman, Gil Bar-Sela, Alexander T. Pearson, Dezheng Huo, Frederick M. Howard, Ron Kimmel, Chen Mayer

**Affiliations:** Taub Faculty of Computer Science, Technion-Israel Institute of Technology, Haifa, Israel; The Institute of Pathology, Carmel Medical Center, Ruth and Bruce Rappaport Faculty of Medicine, Technion-Institute of Technology, Haifa, Israel; Department of Pathology, Sheba Medical Center, Tel Hashomer, Ramat-Gan, Israel; Department of Pathology, Faculty of Medicine, Tel Aviv University, Tel Aviv, Israel; Department of Pathology, Emek Medical Center, Afula, Israel; Department of Oncology, Emek Medical Center, Afula, Israel; Technion Integrated Cancer Center, Faculty of Medicine, Technion-Israel Institute of Technology, Haifa, Israel; Department of Medicine, University of Chicago, Chicago, Illinois, USA; Department of Public Health Sciences, University of Chicago, Chicago, IL, USA; Faculty of Electrical and Computer Engineering, Technion-Israel Institute of Technology, Haifa, Israel

## Abstract

The OncotypeDX 21-gene assay is a widely adopted tool for estimating recurrence risk and informing chemotherapy decisions in early-stage, hormone receptor-positive, HER2-negative breast cancer. Although informative, its high cost and long turnaround time limit accessibility and delay treatment in low- and middle-income countries, creating a need for alternative solutions. This study presents a deep learning-based approach for predicting OncotypeDX recurrence scores directly from hematoxylin and eosin-stained whole slide images. Our approach leverages a deep learning foundation model pre-trained on 171,189 slides via self-supervised learning, which is fine-tuned for our task. The model was developed and validated using five independent cohorts, out of which three are external. On the two external cohorts that include OncotypeDX scores, the model achieved an AUC of 0.825 and 0.817, and identified 21.9% and 25.1% of the patients as low-risk with sensitivity of 0.97 and 0.95 and negative predictive value of 0.97 and 0.96, showing strong generalizability despite variations in staining protocols and imaging devices. Kaplan-Meier analysis demonstrated that patients classified as low-risk by the model had a significantly better prognosis than those classified as high-risk, with a hazard ratio of 4.1 (P<0.001) and 2.0 (P<0.01) on the two external cohorts that include patient outcomes. This artificial intelligence-driven solution offers a rapid, cost-effective, and scalable alternative to genomic testing, with the potential to enhance personalized treatment planning, especially in resource-constrained settings.

## Introduction

Breast cancer remains one of the most prevalent malignancies worldwide, with an estimated 2.3 million new cases diagnosed annually^1^. The management of breast cancer has evolved considerably in recent years, shifting towards more personalized treatment approaches. Central to this paradigm shift is the use of genomic assays, which provide crucial information about tumor biology and help guide treatment decisions, particularly regarding the necessity of adjuvant chemotherapy. OncotypeDX, a 21-gene expression assay, has become a standard tool in clinical practice for early-stage, hormone receptor-positive, human epidermal growth factor receptor 2 (HER2) negative breast cancers^2^. Utilizing RT-PCR gene expression profiling, this test generates a recurrence score (RS) that estimates the risk of distant recurrence. Patients classified as high-risk (RS≥26) were shown to benefit from chemotherapy, whereas no chemotherapy benefit was observed for low-risk (RS<11) and intermediate-risk (11≤RS<26) patients^3,4^. While OncotypeDX offers substantial clinical benefits, it is associated with significant costs and turnaround time, which can delay treatment initiation and limit accessibility in resource-constrained settings.

In recent years, the field of digital pathology has witnessed remarkable advancements, particularly in the application of artificial intelligence (AI) to hematoxylin and eosin (H&E)-stained whole-slide images (WSIs)^5–10^. These developments have opened new avenues for extracting valuable molecular and prognostic information from routine histopathological slides^11–22^.

Several previous studies have explored AI-based methods for predicting OncotypeDX scores, often combining histopathology with clinical or textual data^23–26^. However, these approaches have generally shown low generalizability to external cohorts and included relatively few or small-sized external datasets, highlighting the need for robust models that generalize well across diverse clinical environments.

Self-supervised learning (SSL) enables the extraction of meaningful features from large amounts of unlabeled data, which is especially valuable in pathology, where labeled data is limited and expensive to obtain.

Foundation models, trained using SSL, make use of especially large and diverse datasets across institutions, improving their ability to generalize. This is particularly important in clinical practice, where variability in slide preparation, staining, and imaging protocols can affect performance.

Here, we develop a deep learning model for predicting the OncotypeDX RS from H&E-stained WSIs. Our study offers two main contributions. First, we leverage a foundation model trained via SSL on 171,189 whole-slide images. This model enables us to develop a robust system that is able to generalize to different cohorts despite variations such as different slide preparation or staining protocols. Second, the foundation model is tuned and validated to predict the OncotypeDX RS using diverse data; our data collection consists of 5546 H&E slides from 4,227 patients across five independent cohorts, three of which are external, representing diverse data used to validate the model in a real-world setting. Our results suggest that the proposed model enables immediate and accurate risk stratification. In resource-limited settings where genomic testing is often unavailable, this approach could help deliver personalized treatment decisions, making precision oncology more accessible and potentially improving outcomes for a large number of breast cancer patients worldwide.

## Methods

### Datasets Used

This study utilized H&E-stained WSIs of breast cancer tumors collected from five medical centers **(Table 1)**: Carmel Medical Center (1022 slides from 569 patients), Haemek Medical Center (202 slides from 158 patients), Sheba Medical Center (688 slides from 433 patients), Australian Breast Cancer Tissue Bank (ABCTB) (2961 slides from 2503 patients) and the University of Chicago Medical Center (UCMC) (601 slides from 511 patients). OncotypeDX scores were available for the Carmel, Haemek, Sheba, and UCMC datasets, while survival data was available for the UCMC and ABCTB cohorts. Inclusion and exclusion criteria: From all cohorts, only patients with hormone receptor-positive, HER2-negative breast cancer were included. For Carmel, Haemek, Sheba, and UCMC, only patients with OncotypeDX scores were included. Slides that had physical scan issues or less than 100 available tissue tiles (see Slide segmentation and tilling) were also excluded. Other than the exclusions described above, no further data curation was done. Additional clinical and dataset-specific details are provided in **Supplementary Table 1**.

**Table 1:** Distribution of patients across the different datasets used in development and evaluation.

| Dataset: | Year of diagnosis | Total collected patients (slides) | Included in analysis patients (slides) | Available OncotypeDX data | Available survival data |
| --- | --- | --- | --- | --- | --- |
| Carmel train | 2015-2022 | 427 (767) | 425 (762) | Yes | No |
| Carmel test | 2015-2022 | 142 (255) | 140 (251) | Yes | No |
| Haemek train | 2014-2022 | 135 (176) | 133 (173) | Yes | No |
| Haemek test | 2014-2022 | 23 (26) | 23 (26) | Yes | No |
| Sheba | 2014-2020 | 437 (697) | 427 (676) | Yes | No |
| UCMC | 2006-2020 | 511 (601) | 490 (568) | Yes | Yes |
| ABCTB | 2006-2015 | 2552 (3024) | 1762 (2080) | No | Yes |
| Total |  | 4227 (5546) | 3400 (4536) |  |  |

*Summary of the five cohorts used for development and validation in the study, three of which are from Israel (Carmel, Haemek, Sheba), one from the United States (UCMC), and one from Australia (ABCTB). We present the number of patients and slides available, the number included in the analysis, and whether or not OncotypeDX and survival data were available.*

### Slide segmentation and tilling

During preprocessing, we employed Otsu’s thresholding method^27^ to distinguish between the tissue and background regions of the WSIs. Following the thresholding, we divided the tissue regions into non-overlapping tiles measuring 256×256 at a magnification of 0.5 microns per pixel (MPP) across all cohorts.

### The proposed model

Our model makes use of the Prov-GigaPath foundation model, which was pre-trained on 171,189 whole-slide images^28^. Using a fixed transformer-based tile encoder, we generated a feature vector containing 1536 features per tile image. After producing a feature vector for all the tiles in a given slide, we fine-tuned a transformer-based slide encoder to predict the slide-level OncotypeDX RS. Both the slide and tile encoders were a part of the default pre-trained foundation model. During training, we used a mean squared error loss, the Adamw optimizer with an initial learning rate of 2.5e-4, and a cosine learning rate scheduler. The model was trained for 5 epochs with a batch size of 32.

### Model evaluation and statistical analysis

The Carmel and Haemek datasets were combined into an internal cohort used to develop the models. This internal cohort was split at the patient level, with 75% of patients assigned to the training set and 25% to the test set. The training set was further divided into 5 training folds. Slides from Sheba, ABCTB, and UCMC were used for external validation and survival analysis. Five-fold cross-validation was used to compute the predicted scores, where each model was trained on four of the five folds from the Carmel and Haemek training sets. When evaluating the model on the test sets, all five models produced a score for a given slide, and the scores were then averaged to produce a final slide score. In case of multiple slides per patient, the scores from the different slides were averaged to generate a score at the patient level. The performance metrics used for evaluation were AUC, area under the precision-recall curve (AUPRC), specificity, sensitivity, negative predictive value (NPV), and positive predictive value (PPV). The bootstrap technique with 1000 resampling iterations was used for all metrics to generate 95% confidence intervals (CIs) at the patient level. For survival analysis, Kaplan-Meier estimation was performed, and accuracy was evaluated by measuring a hazard ratio (HR) using Cox regression and P-value using the log-rank test.

### Incorporation of clinical data

To test the potential benefit of including clinical variables, we combined the model-predicted scores with clinical data. A linear classifier was trained using the predicted scores and clinical features as inputs to predict the OncotypeDX score. The clinical features included: Ki67 expression, tumor grade, tumor size in cm, age at diagnosis, ER IHC score, PR IHC score, and HER2 class. The classifier was trained using ordinary least squares, and feature importance was evaluated by examining the normalized coefficients of the classifier. This experiment was done using the Sheba cohort because it had the entire set of clinical variables.

### Score calibration with histogram matching

While the distribution of ground-truth OncotypeDX scores in the different cohorts is relatively similar, the distribution of model-predicted scores presents some variation between cohorts, which caused an issue in some analyses. For example, when using a high sensitivity cut-off threshold to discriminate between low and high-risk patients (see Evaluating the model’s clinical utility), 22% of the patients in the internal cohort would be classified as low-risk. In comparison, 18% and 0.6% of the patients would be classified as low-risk in the UCMC and Sheba cohorts, respectively. To account for this variation, we applied histogram matching between the ground-truth OncotypeDX scores and the model-predicted scores for model calibration. To calibrate for each external cohort, we randomly sampled 100 patients from this cohort and used their data to generate a mapping function. The resulting mapping function from the histogram matching process was then applied to the model’s scores. The ground-truth, as well as model-predicted score distributions, is presented in **Supplementary Fig.1**. Since ABCTB does not have ground-truth OncotypeDX data, we used patients from the internal cohort, which included the Carmel and Haemek datasets. This calibration was relevant for the survival analysis Kaplan-Meier estimation, for evaluating NPV and sensitivity when stratifying patients using a fixed threshold, and for the evaluation of the importance of clinical features when generating binary predictions based on the model scores. In those analyses, the randomly selected patients used to generate the mapping function were excluded. The rest of the presented analyses, which included model performance evaluation by computing AUC, were agnostic to the calibration, as it does not affect ranking-based metrics.

## Results

### Model evaluation on the internal and external test sets

We trained and validated our model using the training set and test set of the combined internal cohorts (Carmel and Haemek). The model achieved an AUC of 0.794 (95% CI: 0.672–0.897) for predicting high OncotypeDX risk (RS≥26) on the combined internal test set (**Fig. 1a** and **Table 2**). This shows that the model is capable of separating patients into low and high genomic risk groups with high reliability. Next, we validated the model on the Sheba and UCMC external cohorts, which had OncotypeDX RS data. The model achieved an AUC of 0.817 (95% CI: 0.77–0.868) on the Sheba cohort (**Fig. 1b)** and an AUC of 0.825 (95% CI: 0.771–0.837) on the UCMC cohort (**Fig. 1c)**, indicating that the model generalizes well to external cohorts. The model achieved a Pearson correlation coefficient of 0.605 and a Spearman correlation coefficient of 0.547 on the Sheba cohort. On the UCMC cohort, the model achieved a Pearson correlation coefficient of 0.519 and a Spearman correlation coefficient of 0.481.

**Table 2:**
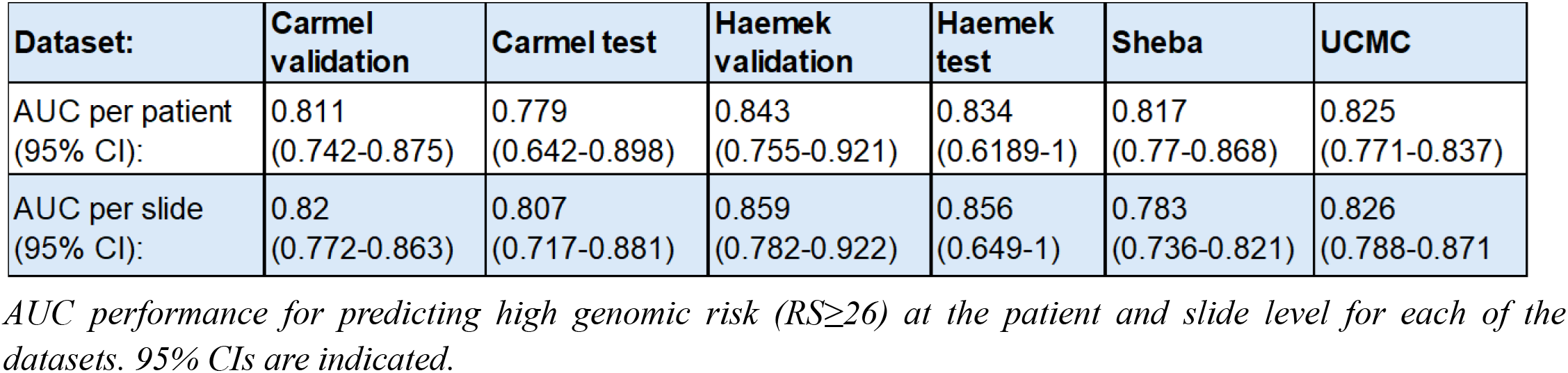
Model AUC performance across the datasets for predicting high genomic risk.

| **Dataset:** | **Carmel validation** | **Carmel test** | **Haemek validation** | **Haemek test** | **Sheba** | **UCMC** |
| --- | --- | --- | --- | --- | --- | --- |
| AUC per patient<br>(95% CI): | 0.811<br>(0.742-0.875) | 0.779<br>(0.642-0.898) | 0.843<br>(0.755-0.921) | 0.834<br>(0.6189-1) | 0.817<br>(0.77-0.868) | 0.825<br>(0.771-0.837) |
| AUC per slide<br>(95% CI): | 0.82<br>(0.772-0.863) | 0.807<br>(0.717-0.881) | 0.859<br>(0.782-0.922) | 0.856<br>(0.649-1) | 0.783<br>(0.736-0.821) | 0.826<br>(0.788-0.871) |

*AUC performance for predicting high genomic risk ( $RS \geq 26$ ) at the patient and slide level for each of the datasets. 95% CIs are indicated.*

**Figure 1:**
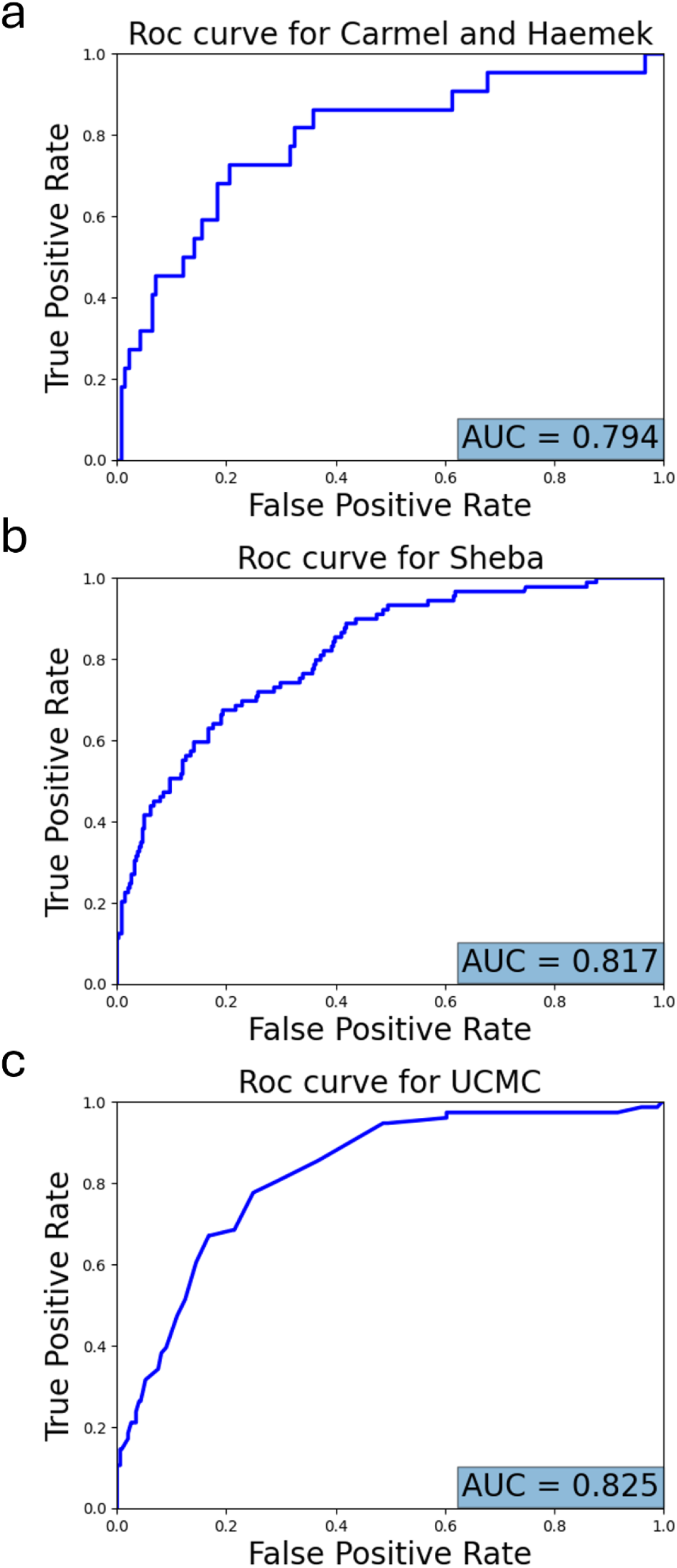
Model AUC performance. ROC curves obtained by the model for predicting high genomic risk (RS≥26) for: **(a)** the test set of the internal cohorts (Carmel and Haemek). **(b)** Sheba. **(c)** UCMC. AUC value for each graph is specified at the bottom.

### Evaluation of discrimination ability versus cut-off threshold

The initial validation studies, NSABP B-20 and SWOG-8814, demonstrated significant chemotherapy benefit in patients with RS≥31^29,30^. A reanalysis of the B-20 cohort indicated that RS≥26 provided a more appropriate threshold for the prediction of chemotherapy benefit^31,32^, which was later used in the TAILORx study^3^. Yet, the optimal OncotypeDX threshold to determine chemotherapy benefit remains uncertain^33^. We next evaluated the model’s ability to discriminate low and high-genomic risk patients for cut-off thresholds other than 26. Namely, for each threshold ***t***, we computed the AUC and balanced AUPRC performance of the model for predicting OncotypeDX RS≥***t*** in the internal and external cohorts (**Supplementary Fig. 2**). The analysis shows that the AUC and balanced AUPRC performance steadily rose with respect to ***t***, indicating that the model’s ability to distinguish between low and high genomic risk improved with higher cut-off thresholds. Both AUC and balanced AUPRC were computed to evaluate both the actual model performance and the effects of potential imbalances between the positive and negative classes on model performance. When computing AUPRC, we chose to balance the data through subsampling the majority class to ensure a fair comparison of the performance for different thresholds. Interestingly, this indicates that cut-off thresholds higher than 26 exhibited more distinctive morphological characteristics.

### Evaluating the model’s clinical utility

We next aimed to assess the clinical utility of the model. To this end, we evaluated the model’s specificity, sensitivity, NPV, and PPV on the Sheba and UCMC cohorts for different cut-off thresholds **(Fig. 2)**. Notably, when using a high sensitivity threshold, computed in a manner used in previous studies^34^ to achieve sensitivity=0.95 on the internal cohorts, the model could identify 21.9% and 25.1% of the patients who had a low genomic score, with sensitivity of 0.97 and 0.95 and NPV of 0.97 and 0.96, showing strong performance in identifying low genomic risk patients (OncotypeDX < 26). This demonstrates the model’s potential to spare unnecessary chemotherapy for many patients, which may be particularly relevant in countries without access to genomic assays.

**Figure 2:**
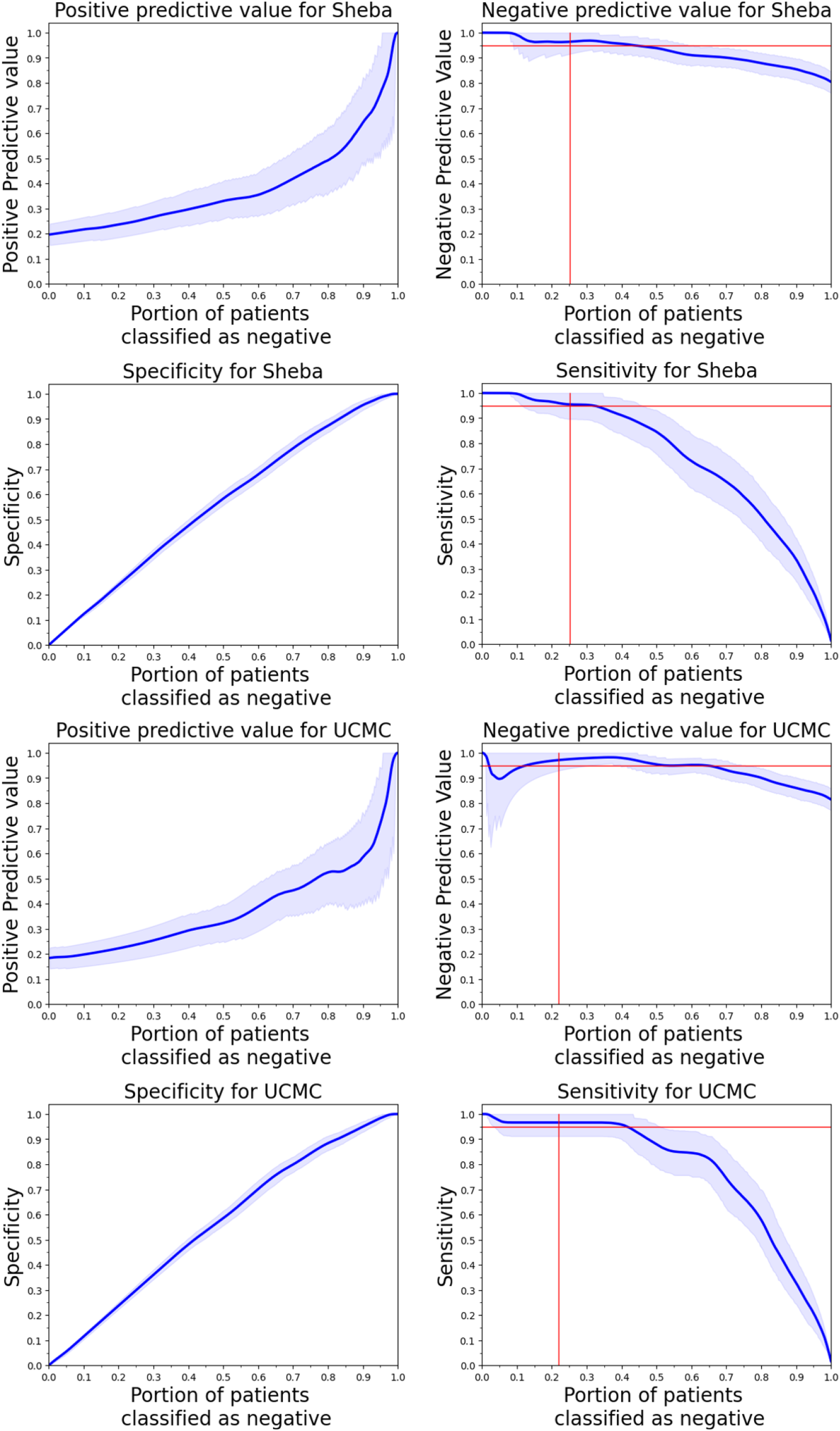
Clinical utility evaluation on the external cohorts. PPV, NPV, specificity, and sensitivity per portion of patients classified as negative for the external test sets (Sheba and UCMC) (dark blue line) and 95% CI (light blue interval). Red lines extended at the low-risk cutoff threshold for each cohort and the 95% NPV and sensitivity points.

### Incorporation of clinical data

We next evaluated the potential benefit of including clinical variables in the training process. To this end, we trained a linear classifier to predict the OncotypeDX scores from both H&E and clinical variables on the Sheba cohort, which had sufficient clinical variables available (Methods). We then conducted a feature importance analysis, which revealed that the model-predicted score was the most significant predictor, followed by the PR expression as the second most important feature **(Fig. 3)**.

**Figure 3:**
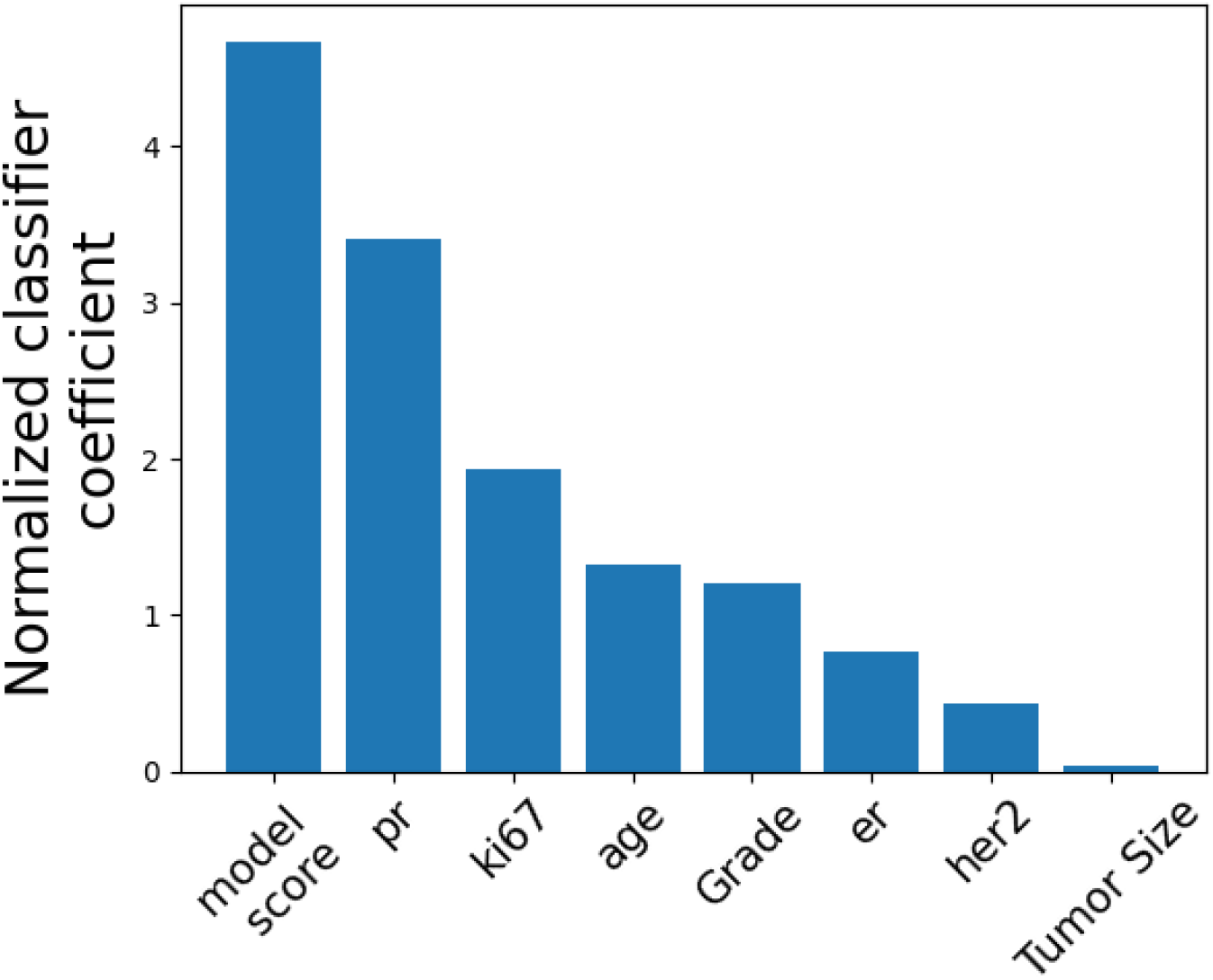
Feature importance by coefficient score. Feature importance measured by normalized coefficient values for the linear classifier trained on the Clinical features and slide model scores on the Sheba cohort.

### Survival analysis

We next evaluated the model’s ability to estimate patient prognosis. We stratified the patients in the ABCTB and UCMC cohorts into low and high-risk groups based on the calibrated model scores **(Fig. 4)**. On UCMC, our endpoint was recurrence-free interval, achieving HR=4.1 (P<0.001). On ABCTB, our endpoint was breast cancer-specific survival because no recurrence data were available in this cohort, achieving HR=2.0 (P < 0.01). This analysis shows that the model could stratify patients into low and high risk.

**Figure 4:**
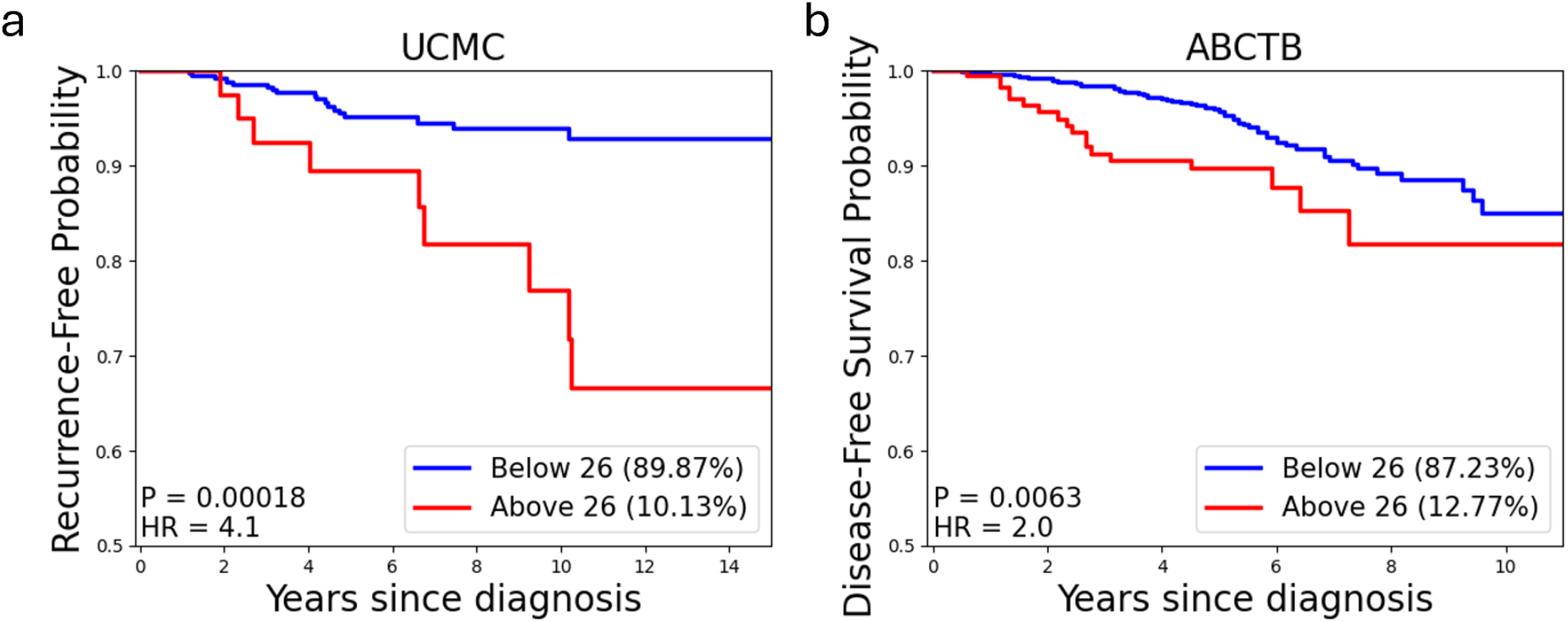
Survival analysis for UCMC and ABCTB. Kaplan-Meier analysis for: **(a)** Recurrence-free interval for UCMC. **(b)** Breast Cancer-specific survival for ABCTB. For each cohort, patients classified as low-risk by the model are represented by a blue curve, and patients classified as high-risk are represented by a red curve. The portion of patients in each risk group, as well as P values and HR, are indicated at the bottom.

## Discussion

In this study, we have developed a system aimed at predicting the OncotypeDX score in breast cancer patients directly from H&E-stained tissue slides. OncotypeDX is a widely used genomic assay that quantifies the likelihood of recurrence in breast cancer patients, helping clinicians make informed treatment decisions. However, its high cost, time-consuming nature, and the logistical challenges of sending specimens to specialized laboratories for testing limit its accessibility and turnaround time in some clinical settings. The test itself can take several weeks to return results, which can delay treatment decisions. By leveraging AI, particularly deep learning models, it is possible to approximate OncotypeDX scores based solely on histopathological features visible in H&E slides, offering the potential for a less invasive, more cost-effective, and faster alternative.

Our AI-driven model demonstrated a strong correlation with the OncotypeDX score, suggesting that it is able to capture critical histopathological features linked to tumor biology that influence recurrence risk. Histologically, H&E-stained slides provide a wealth of information about the structural and cellular composition of tumors, including features such as nuclear atypia, mitotic activity, stromal characteristics, and the extent of immune infiltration. These features are known to reflect underlying biological processes, such as proliferation, genetic instability, and tumor microenvironment interactions, which are directly linked to recurrence risk and response to therapy. While traditional pathology relies on human interpretation to identify these features, AI excels at detecting subtle, complex patterns and relationships within the data that may not be readily apparent to the human eye.

A key innovation in our approach is the use of a foundation model—Prov-GigaPath— which was pretrained using SSL. This deep learning model is pre-trained on a large and diverse pathology dataset and fine-tuned for the specific task of predicting OncotypeDX scores from H&E-stained slides. Foundation models have demonstrated effectiveness across various domains, including natural language processing, computer vision, and medical imaging, due to their ability to learn transferable representations from broad, unlabeled data^28,35,36^. This approach is particularly advantageous in clinical contexts, where curated annotations are costly, class distributions are imbalanced, and privacy constraints limit data sharing.

By starting from a broadly pre-trained model, we were able to reduce reliance on task-specific data while achieving strong predictive performance. Crucially, the model’s pretraining on heterogeneous pathology data enhances its ability to generalize across institutional, demographic, and technical variability, such as differences in staining protocols or imaging devices, supporting its robustness in real-world applications. This generalizability is a key benefit of the foundation model paradigm and underpins its suitability for scalable clinical deployment.

Previous studies have shown the potential of predicting OncotypeDX from H&E-stained slides^23,24^. However, these studies exhibited limitations in generalizability and were constrained by relatively small training datasets and narrower validation cohorts. Boehm et al. reported strong internal performance with their image-only model (AUC 0.85). However, they showed a decline in AUC on their external cohorts (0.81 and 0.80), which included 1027 patients^23^. Similarly, Goyal et al. also demonstrated a decline in performance between the internal cohort (AUC 0.91) and the external cohort (AUC 0.84), which consisted of 405 patients^24^. In contrast, our model maintained consistent AUCs across internal (0.794) and external (0.817 and 0.825) cohorts, demonstrating the robustness of the foundation model approach. Additionally, we utilized a foundation model trained on 171,189 slides, an order of magnitude larger than foundation models used in earlier works, enabling strong generalization across diverse data sources. Our study included 727 patients from two internal cohorts and 3,500 patients from three external cohorts, encompassing substantial variation in geographic origin, patient demographics, and slide preparation protocols across multiple laboratories. These findings underscore the broad applicability of our approach, particularly in heterogeneous real-world settings.

Despite the promising results, several limitations of this study must be considered. First, when we tested the inclusion of additional clinical features on the Sheba cohort, we observed that other features also held importance for prediction. Additionally, several different studies utilized a multimodal approach and showed promising results^23,37^. However, since these features were unavailable in the rest of our dataset, a broader assessment was not possible. This highlights the potential of improving our system’s performance with a multimodal approach, which future studies could explore to enhance breast cancer outcome prediction.

Another limitation is the interpretability of the AI model. While deep learning algorithms can achieve high predictive accuracy, their decision-making processes are often opaque, making it difficult for clinicians to understand how specific features in the H&E slides are contributing to the final prediction. This “black box” nature of AI models can be a barrier to their clinical adoption, as clinicians may hesitate to rely on predictions that lack transparency. Future research efforts should focus on improving the explainability of these AI models, enabling clinicians to gain more insight into the factors influencing the prediction and thus fostering greater trust in the technology.

In conclusion, our work introduces a model that can reliably predict OncotypeDX scores from H&E-stained breast cancer tissue slides, offering a promising, quick, and cost-effective addition to traditional genomic assays. By enabling faster and more efficient risk stratification, this approach could significantly improve personalized treatment decisions and expand access to precision medicine for a broader range of patients.

## Ethical approvals for the datasets

All the datasets used in this study were collected in compliance with the Helsinki Declaration, institutional policies, and Data Use Agreements. All the data was de-identified. For each dataset, the respective Institutional Review Board granted ethical approval. For the ABCTB dataset, ethical and scientific approval was received according to the Australian Breast Cancer Tissue Bank access policy.

## Data availability

The ABCTB dataset, accessible from the Australian Breast Cancer Tissue Bank, is subject to ethical and scientific approvals as described in their access policy: https://nsw.biobanking.org/biobanks/view/7. The remaining data collected from medical centers are not available for public access due to privacy and ethical considerations, in alignment with the Helsinki agreements and institutional policies. Interested researchers may request access directly from the respective institutions: Carmel data from the Carmel Medical Center, Israel. Haemek data from the Haemek Medical Center, Israel. Sheba data from Sheba Medical Center, Israel. UCMC data from the University of Chicago Medical Center, USA.

## Code availability

The source code used in this study has been archived in Zenodo^38^ and can be accessed at the following link: https://github.com/shachar5020/TransformerWSI4OncoDXPrediction. Statistical analysis, preprocessing, tuning of the models, and inference were done using Python 3.9.18 and Pytorch library version 2.0.0. Additional Python libraries used for database management, graphical plotting, scientific calculations, and other tasks include Numpy v.1.26.4, Pandas v.2.2.2, Scipy v.1.13.1, and Openslide v.1.3.1.

## Acknowledgments

This research was supported by the Israel Innovation Authority—Kamin 69997 (R.K. and G.S.), the Zimin Institute for Artificial Intelligence Solutions in Healthcare grant (R.K. and G.S.), and the Israel Precision Medicine Partnership program (IPMP) grant 3864/21 (R.K. and G.S.). We would like to thank Karin Stoliar for helping with the data acquisition and quality assurance, Hen Davidov for supporting the deep learning experiments, and Liat Dizengoff for managing the Helsinki approvals in Carmel Medical Center.

---

**Supplementary Table 1:**
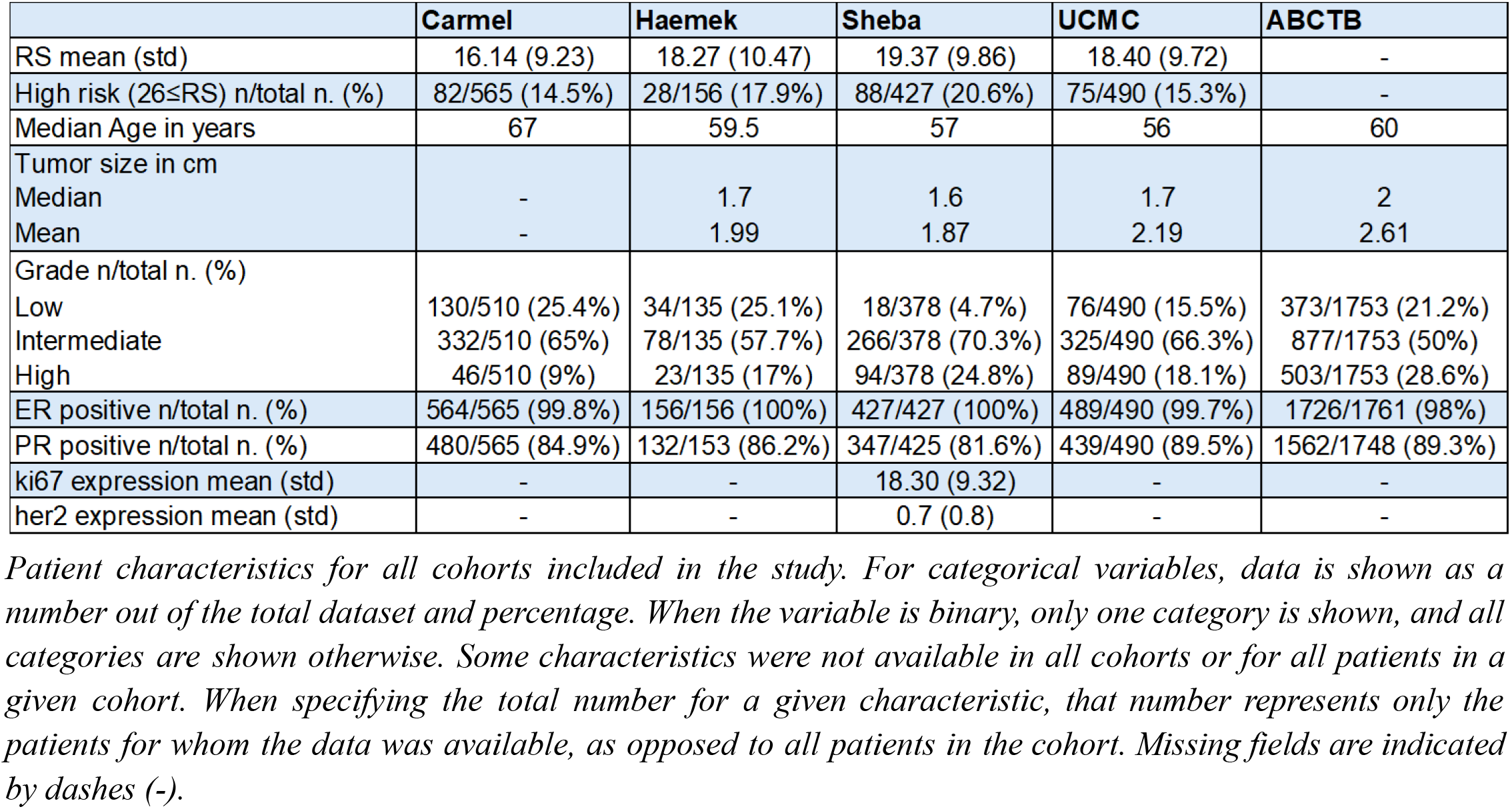
Clinical characteristics of the data.

**Supplementary Figure 1:**
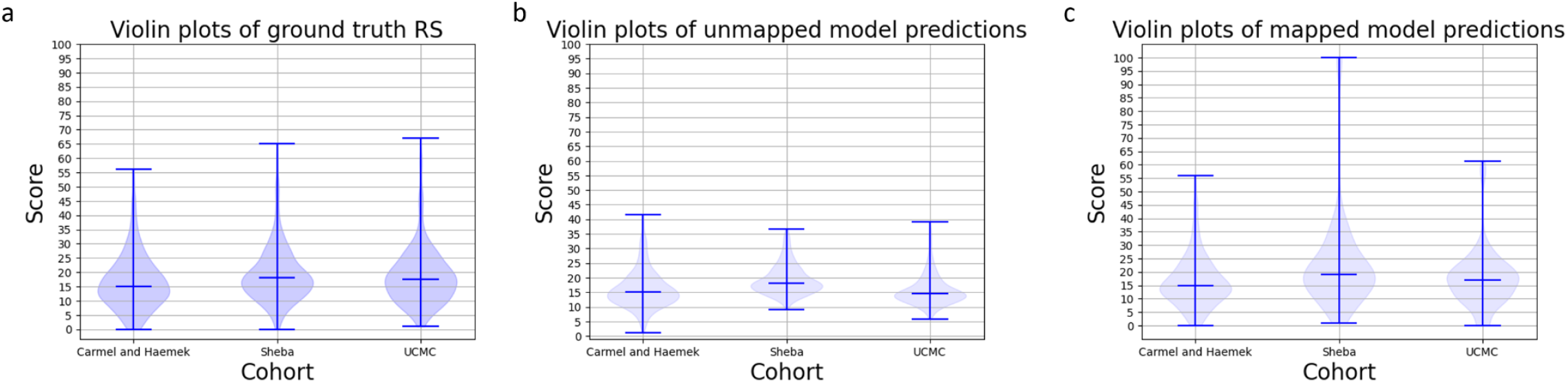
Violin plots representing the RS distribution in the different cohorts. Violin plots of the ground-truth RS **(a)**, unmapped model predictions **(b)**, and the model predictions mapped via histogram matching (see methods) **(c)**. Blue horizontal lines represent the minimal, maximal, and median score for each dataset, and grey horizontal lines are added for ease of comparison.

**Supplementary Figure 2:**
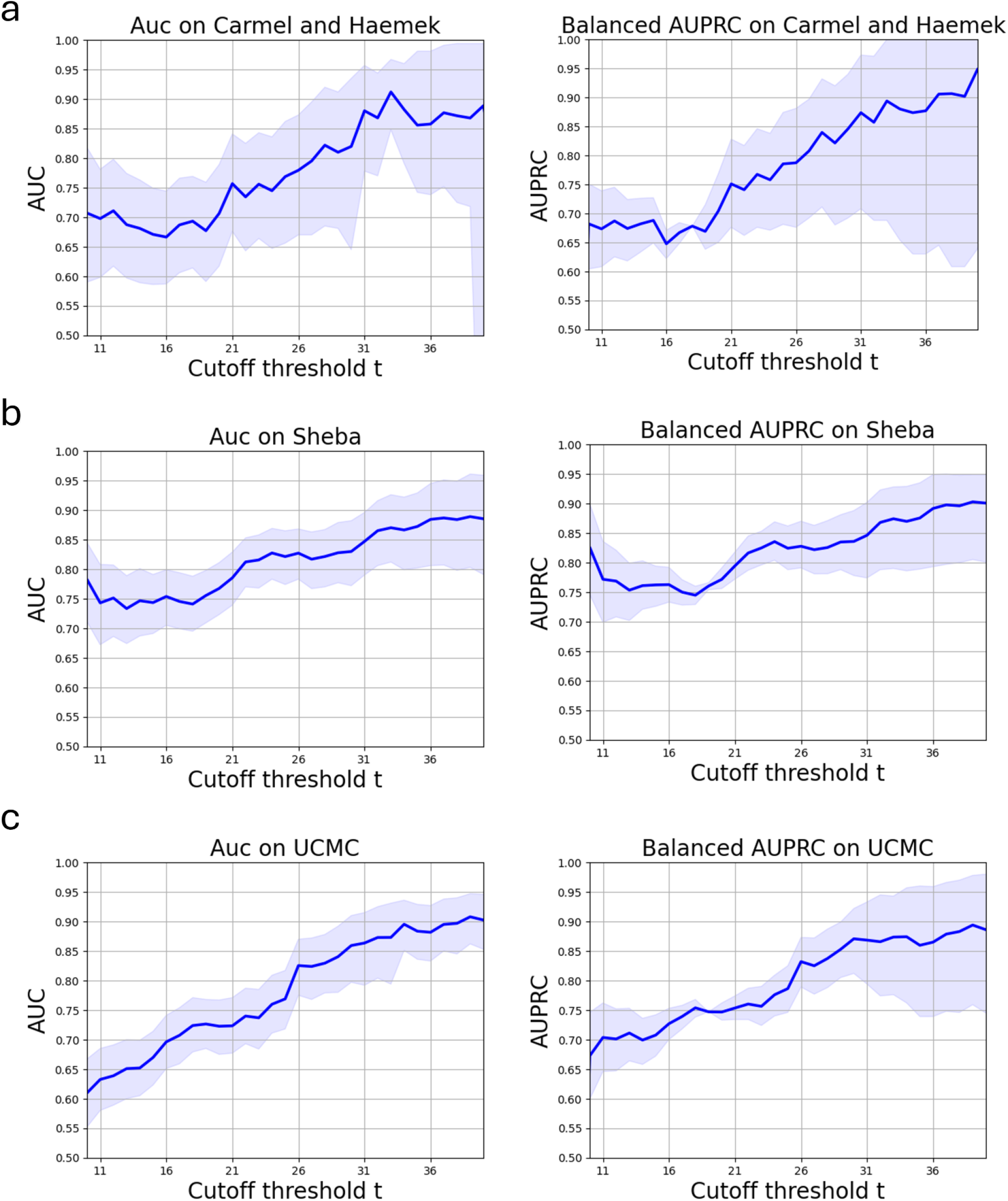
Model AUC as a function of high-risk cutoff threshold. AUC and balanced AUPRC scores obtained by the model for predicting RS≥**t**, using different cutoff thresholds **t** (dark blue line) for: **(a)** the test set of the internal cohorts (Carmel and Haemek). **(b)** Sheba. **(c)** UCMC. The 95% CIs are displayed as a light blue interval.

