## Supplementary tables and figures for "Prediction of OncotypeDX recurrence score using H&E stained WSI images"

**Supplementary Table 1: Clinical characteristics of the data**

| | Carmel | Haemek | Sheba | UCMC | ABCTB |
|---|---|---|---|---|---|
| RS mean (std) | 16.14 (9.23) | 18.27 (10.47) | 19.37 (9.86) | 18.40 (9.72) | - |
| High risk (26≤RS) n/total n. (%) | 82/565 (14.5%) | 28/156 (17.9%) | 88/427 (20.6%) | 75/490 (15.3%) | - |
| Median Age in years | 67 | 59.5 | 57 | 56 | 60 |
| Tumor size in cm<br>Median<br>Mean | <br>-<br>- | <br>1.7<br>1.99 | <br>1.6<br>1.87 | <br>1.7<br>2.19 | <br>2<br>2.61 |
| Grade n/total n. (%)<br>Low<br>Intermediate<br>High | <br>130/510 (25.4%)<br>332/510 (65%)<br>46/510 (9%) | <br>34/135 (25.1%)<br>78/135 (57.7%)<br>23/135 (17%) | <br>18/378 (4.7%)<br>266/378 (70.3%)<br>94/378 (24.8%) | <br>76/490 (15.5%)<br>325/490 (66.3%)<br>89/490 (18.1%) | <br>373/1753 (21.2%)<br>877/1753 (50%)<br>503/1753 (28.6%) |
| ER positive n/total n. (%) | 564/565 (99.8%) | 156/156 (100%) | 427/427 (100%) | 489/490 (99.7%) | 1726/1761 (98%) |
| PR positive n/total n. (%) | 480/565 (84.9%) | 132/153 (86.2%) | 347/425 (81.6%) | 439/490 (89.5%) | 1562/1748 (89.3%) |
| ki67 expression mean (std) | - | - | 18.30 (9.32) | - | - |
| her2 expression mean (std) | - | - | 0.7 (0.8) | - | - |

*Patient characteristics for all cohorts included in the study. For categorical variables, data is shown as a number out of the total dataset and percentage. When the variable is binary, only one category is shown, and all categories are shown otherwise. Some characteristics were not available in all cohorts or for all patients in a given cohort. When specifying the total number for a given characteristic, that number represents only the patients for whom the data was available, as opposed to all patients in the cohort. Missing fields are indicated by dashes (-).*

**Supplementary Figure 1: Violin plots representing the RS distribution in the different cohorts**

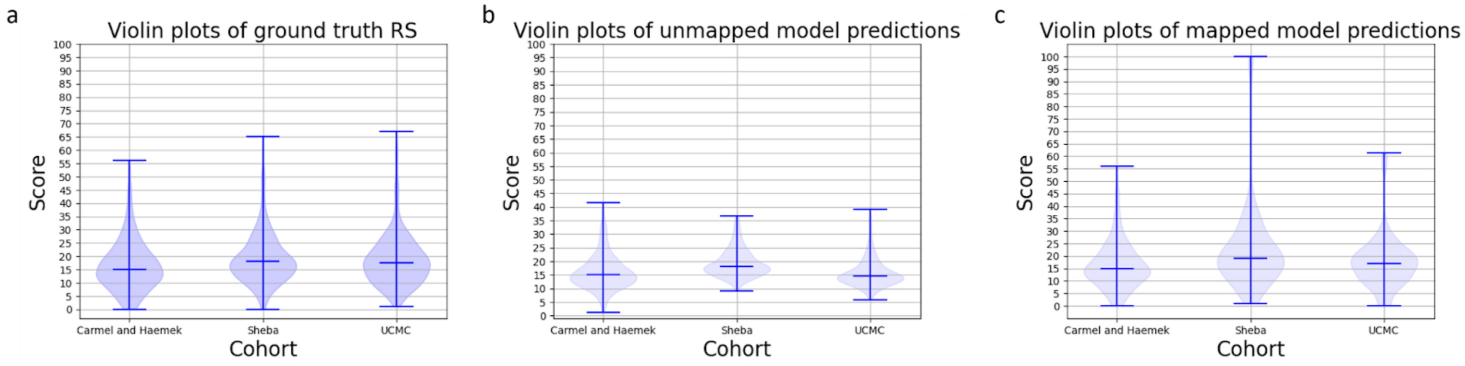


*Violin plots of the ground-truth RS **(a)**, unmapped model predictions **(b)**, and the model predictions mapped via histogram matching (see methods) **(c)**. Blue horizontal lines represent the minimal, maximal, and median score for each dataset, and grey horizontal lines are added for ease of comparison.*

**Supplementary Figure 2: Model AUC as a function of high-risk cutoff threshold**

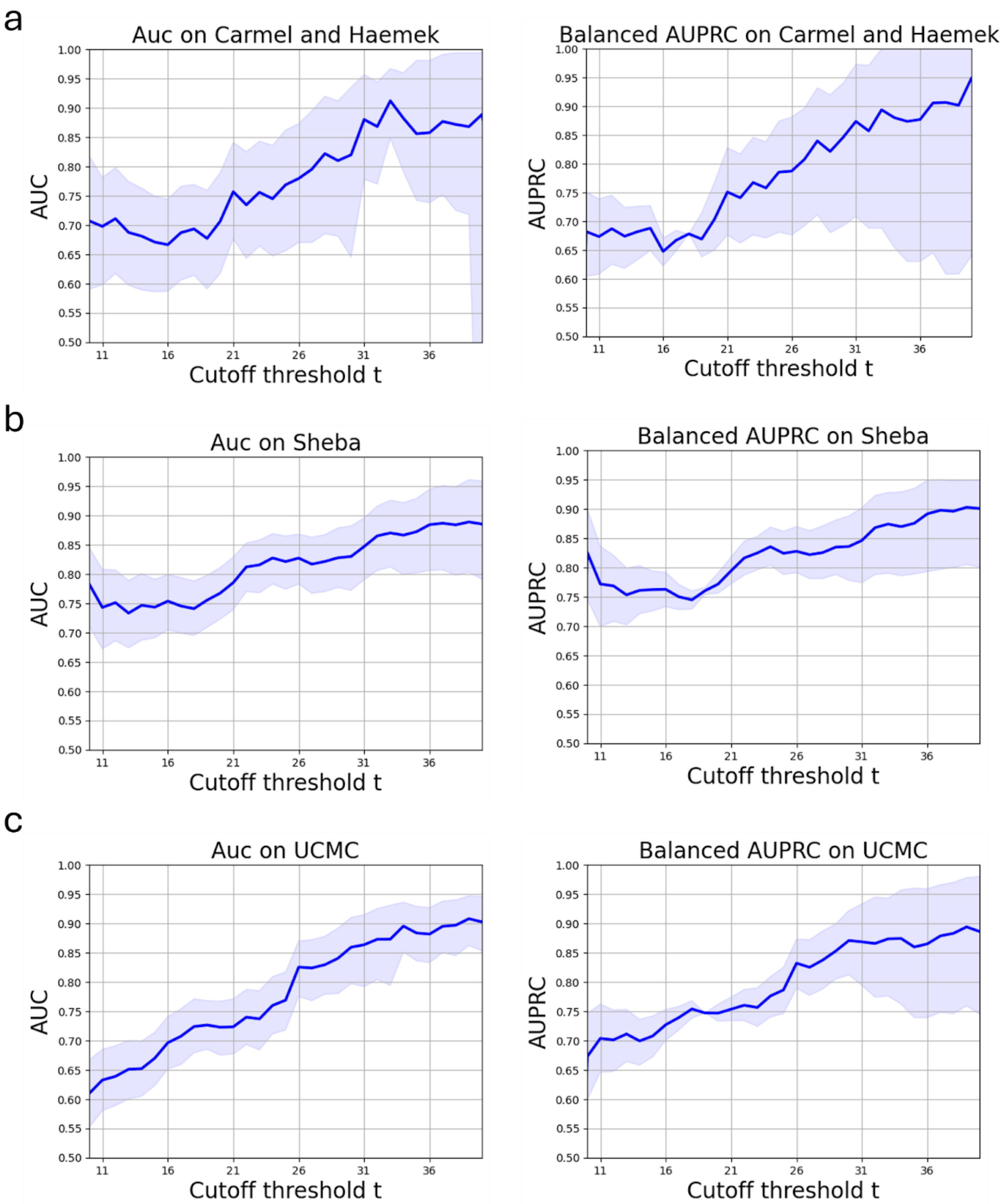


*AUC and balanced AUPRC scores obtained by the model for predicting RS≥**t**, using different cutoff thresholds **t** (dark blue line) for: **(a)** the test set of the internal cohorts (Carmel and Haemek). **(b)** Sheba. **(c)** UCMC. The 95% CIs are displayed as a light blue interval.*
